# Predictors of QT Interval Prolongation in Critically-ill Patients with SARS-CoV-2 Infection Treated with Hydroxychloroquine

**DOI:** 10.1101/2020.11.26.20239418

**Authors:** Frederico Scuotto, Rogério Marra, Lilian Leite de Almeida, Mariana Santa Rita Soares, Gabriela Kurita Silva, Luiz Carlos Paul, Guilherme Drummond Fenelon Costa, Cláudio Cirenza

## Abstract

**Background:** Hydroxychloroquine (HCQ) has been described as a potential treatment for SARS-CoV-2 infection. However, there are safety concerns regarding its QT interval and pro-arrhythmic effects.

**Objective:** This trial aimed to determine the predictors of QT interval prolongation and pro-arrhythmic effects in patients hospitalized for SARS-CoV-2 infection and receiving HCQ.

**Methods:** We performed a retrospective observational study of 45 critically-ill patients hospitalized because of SARS-CoV-2 infection and treated with 800 mg of HCQ at day 1 and 400 mg on days 2–5. Clinical aspects and outcomes, basal and final corrected QT (QTc) interval, and the incidence of arrhythmias and arrhythmogenic death were observed. Independent predictors of QTc prolongation were identified using multivariable logistic regression analysis. QT interval prolongation was considered substantial at final QTc ≥ 480 ms.

**Results:** The mean age was 60.9 ± 16.67 years and 28 (62.2%) patients were men. Basal QTc was 442 ± 28 ms, and the final QTc interval was 458 ± 34 ms, for a mean QTc interval variation of 15 ± 11 ms. There was no arrhythmogenic death. The need for hemodialysis remained a statistically significant predictor of QT interval enlargement (odds ratio, 10.34; 95% confidence interval, 1.04 – 102.18; p = 0.045).

**Conclusions:** HCQ promotes mild to moderate QT interval prolongation. The risk of QT interval prolongation is higher among patients with acute renal failure requiring hemodialysis.

## Introduction

The urgent need for a treatment response for the COVID-19 pandemic led to the “off-label” use of medications, such as chloroquine and hydroxychloroquine (HCQ), which was supported by in vitro trials^1^ and minor clinical trials,^2^ but remain under evaluation in larger randomized clinical trials. In Brazil, the use of HCQ alone or associated with the macrolide antibiotic azithromycin was designated by the Brazilian Ministry of Health only for severe cases of respiratory failure in the beginning of the COVID-19 pandemic.^3^ However, each of these medications has the potential to lengthen the QT interval on the electrocardiogram (ECG) and lead to severe ventricular arrhythmias, including torsades de pointes (TdP) and polymorphic ventricular tachycardia (VT); furthermore, taken together, there is a risk of synergic effect.^4^

SARS-CoV-2 infection may lead to respiratory and renal failure, prothrombotic status, and myocardial injury, which may precipitate worsening of heart failure, acute myocardial infarction (AMI), and cardiac arrhythmias.^5,6,7^ Hence, the pro-arrhythmic effects of this treatment regimen might be potentially heightened in the critically-ill form of the SARS-CoV-2 infection, requiring proper investigation in this subset of patients. Thus, we conducted an observational retrospective study of 45 patients critically ill with SARS-CoV-2 infection in order to evaluate the safety of the proposed treatment as well as its effect on the QT interval in this specific subset of patients.

## Methods

### Study Design

We conducted an observational retrospective study utilizing data from patients treated with HCQ, as indicated for patients with SARS-CoV-2 infection and severe respiratory failure according to the first protocol designated by the Brazilian Ministry of Health in the beginning of the COVID-19 pandemic.

The study was submitted to the hospital’s ethics committee and approved. Since the study was retrospective and observational, no informed consent was required.

### Patient Selection

Since the beginning of the outbreak until May 20, 2020, 45 patients critically ill with SARS-CoV-2 infection were treated with HCQ at our institution.

The inclusion criteria were age ≥18 years, reverse-transcriptase polymerase chain reaction (RT-PCR)-confirmed SARS-CoV-2 infection, and critically ill clinical condition. The parameters to define the critical condition, indication for HCQ, and drug dosage are summarized in Table 1.

**Table 1.** Brazilian Ministry of Health Guideline for HCQ treatment for SARS-CoV-2 infection

| Clinical condition | Treatment Regimen | Recommendation |
| --- | --- | --- |
| **Inpatient with Severe Form of SarS-CoV-2 Infection**<br>(respiratory rate ≥ 30/min, O<sub>2</sub> Sat ≤93%, PaO<sub>2</sub>/FiO<sub>2</sub> <300, and/or lung injury > 50%) within 24-48 h of admission | **Hydroxychloroquine**<br>400 mg BID – first day<br>400 mg PO – 2<sup>nd</sup> – 5<sup>th</sup> day | Check ECG and QTc interval before the beginning of treatment, as well as the subsequent days of treatment.<br>Major risk of QTc interval prolongation with co-administration of other potential risk medications. |
| **Critically-Ill Patient with SARS-CoV-2 Infection**<br>(respiratory failure, septic shock or multiple organ dysfunction) | **Hydroxychloroquine**<br>400 mg BID – first day<br>400 mg PO – 2<sup>nd</sup> – 5<sup>th</sup> day | Check ECG and QTc interval before the beginning of treatment, as well as the subsequent days of treatment.<br>Major risk of QTc interval prolongation with co-administration of other potential risk medications. |

Exclusion criteria included mild or moderate forms of the SARS-CoV-2 infection or suspected cases of COVID-19 without laboratory confirmation.

### Electrocardiogram Analysis

For the electrocardiogram (ECG) parameters, values from the baseline ECG, including heart rate (HR), QRS duration, and morphology as well as measured and corrected QT (QTc) interval, were collected for each patient before the treatment. The QT interval was manually measured from the beginning of the Q wave to the end of the T wave. QT intervals were measured in Lead II. If the QT interval could not be measured in lead II, leads V5 or V6 were alternatively used. Bazzet’s formula was used to calculate the QTc interval. If the patient presented with left bundle branch block (LBBB), Yankelson’s formula was employed.

The hospital’s evaluation protocol included ECG recordings on the 2^nd^ and 4^th^ days of treatment as well as on the first day after the end of treatment regimen (6^th^ day). For patients in critical condition for whom ECG performance was not feasible, monitor recordings were obtained on a daily basis, with viability to measure the heart rate (HR) and QT interval. The QTc interval was measured and calculated by a certified electrophysiologist. Pre-specified criteria for QTc interval prolongation was defined as a final QTc ≥480 ms.

### Predictor Variables

Patient data, including demographic parameters, comorbidities, use of concomitant medications, and time since the onset of symptoms until patient admission, were analyzed. The clinical characteristics at admittance, taking into account the oxygenation upon arrival and the presence of lung compromise on CT scan, were noted. The in-hospital evolution was also evaluated, including the need for intensive care unit (ICU), mechanical ventilation (MV), hemodialysis, vasoactive drugs, and laboratory testing during the HCQ treatment regimen. Any incidence of aborted cardiac arrest, acute myocardial dysfunction, arrhythmias, arrhythmogenic death, and/or patient mortality was also noted.

Concomitant QT interval prolonging agents during treatment were also noticed. Any premature discontinuation of the treatment regimen due to QT prolongation was extracted from medical records. HCQ was frequently associated with azithromycin, which also might lengthen the QT interval. Because of the high rate of association between these two drugs, azithromycin was analyzed separately from other concomitant QT prolonging drugs.

Five patients could not have their final QTc determined, three due to poor clinical evolution and inability to undergo an ECG or proper cardiac monitor registry, and two as a result of premature discharge. However, these patients were maintained within the report due to their clinical evolution and laboratory data as well as clinical outcomes.

### Statistical Analysis

Data are presented as mean ± SD values for numeric variables with normal distribution, median (interquartile range [IQR]) for numeric variables with non-parametric distribution, and percentage (absolute frequency) for categorical variables. Comparisons of characteristics between patients with and without a final QTc ≥480 ms were made using Student’s t-test, Mann-Whitney, and Chi-Square tests for normal numeric variables, non-parametric numeric variables, and categorical variables, respectively. QTc variation before and after the treatment regimen was assessed using the Shapiro-Wilk test. The relationship among the quartile ranges was compared using Student’ s t-test.

Independent predictors of QTc prolongation were identified using multivariable logistic regression analysis. On univariate analysis, p values ≤0.10 were considered to estimate possible predictors. Subsequently, these predictors were evaluated on multivariate analysis, and p values ≤0.05 were considered statistically significant. Multivariate analysis was used to calculate the hazard ratio (HR) and 95% confidence interval (CI).

Statistical analyses were performed using R Software Version 4 for Mac (R Foundation for Statistical Computing, Auckland, NZ).

## Results

The evolution and baseline characteristics of the 45 patients are displayed in Table 2. The mean age was 60.9 ± 16.67 years-old, and 28 (62.2%) of the patients were men. The most common comorbidity was hypertension (51.1%). Coronary heart disease was present in 13.3% of the patients and diabetes in 28.8% of patients. Two patients (4.44%) presented with end-stage renal disease and chronically required renal substitution therapy with hemodialysis.

**Table 2.** Clinical characteristics of the 45 patients under the treatment regimen

|  |  |
| --- | --- |
| **Age (y)** | 60.9 ± 16.67 |
| **Male** | 28 (62.2) |
| **Hypertension** | 23 (51.1) |
| **Coronary Heart Disease** | 6 (13.3) |
| **Heart Failure** | 2 (4.44) |
| **Diabetes** | 13 (28.8) |
| **Previous arrhythmias** | 3 (6.67) |
| **End-stage renal disease** | 2 (4.44) |
| **Concomitant QT prolonging medication** | 15 (33.3) |
| **Azithromycin in Treatment Regimen** | 37 (82.2) |
| **Beginning of symptoms (days)** | 5.75 ± 3.01 |
| **Oxygen Saturation in admittance (median – IQR)** | 88% (85%-90%) |
| **CT scan compromised** | 45 (100) |
| **ICU need** | 42 (93.3) |
| **Mechanical Ventilation** | 32 (71.2) |
| **Hemodialysis** | 23 (51.1) |
| **Acute Heart Failure** | 3 (6.67) |
| **Vasoactive Drug** | 30 (66.7) |
| **Aborted Cardiac Arrest** | 3 (6.67) |
| **Incidental Arrhythmias** | 7 (15.5) |
| **Arrhythmogenic Death** | 0 |
| **Mortality** | 12 (26.6) |

Continuous variables were expressed as mean ± standard deviation (SD). Categorical variables are presented as n (%). ICU intensive care unit, IQR interquartile range

The time since the onset of symptoms was 5.75 ± 3.01 days, and the time between hospital admittance and the initiation of the treatment regimen was 2.0 (IQR, 1.0–4.0) days. Concomitant medication that could lengthen the QTc interval was present in 33.3% of patients, and included antidepressants, antipsychotics, and antiarrhythmics, such as amiodarone. As for the treatment regimen, 82.2% received HCQ plus azithromycin, while 17.8% received only HCQ.

At admittance, signs of respiratory failure were found in most patients, with a mean oxygen saturation of 88% (IQR, 85–90%). All patients presented with altered CT scans, showing signs of viral pneumonia. Among this patient subset, 93.3% required an ICU stay and 71.2% required mechanical ventilation. Of those admitted to the ICU, 66.7% received vasoactive drug support and 51.1% required renal substitution therapy with hemodialysis. Acute heart failure evolved in 6.67% of the patients and the same percentage presented with non-arrhythmic aborted cardiac arrest (pulseless electrical activity or asystole). The overall mortality rate was 26.6%.

The basal QTc interval was 442 ± 28 ms, and the final QTc interval was 458 ± 34 ms, for a mean QTc interval variation of 15 ± 11 ms. The Shapiro-Wilk test showed normal data distribution (p = 0.7173), and the bilateral t-Student’s test showed a statistical difference between the basal and final QTc intervals (p = 0.018). Among the 40 patients with a final QTc measure, 11 (27.5%) presented with a final QTc interval ≥480 ms, and 4 (10%) presented with QTc interval ≥500 ms. Four patients (10%) showed a Δ QTc ≥60 ms. The QTc interval distribution, density, and variation before and after the treatment regimen are depicted in Figures 1, 2, and 3, respectively.

**Figure 1.**
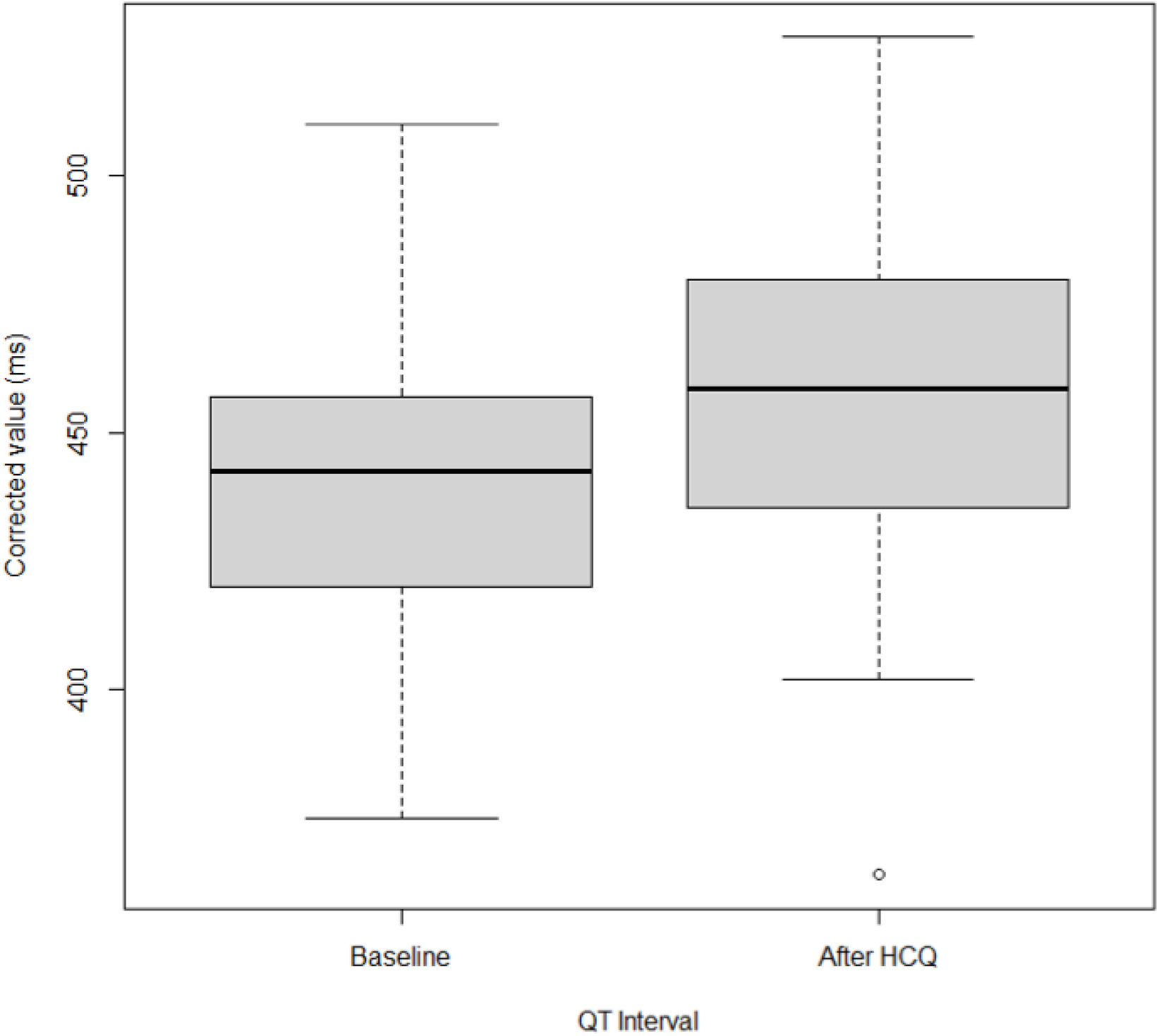
QTc Interval Distribution Before and After Treatment Regimen

**Figure 2.**
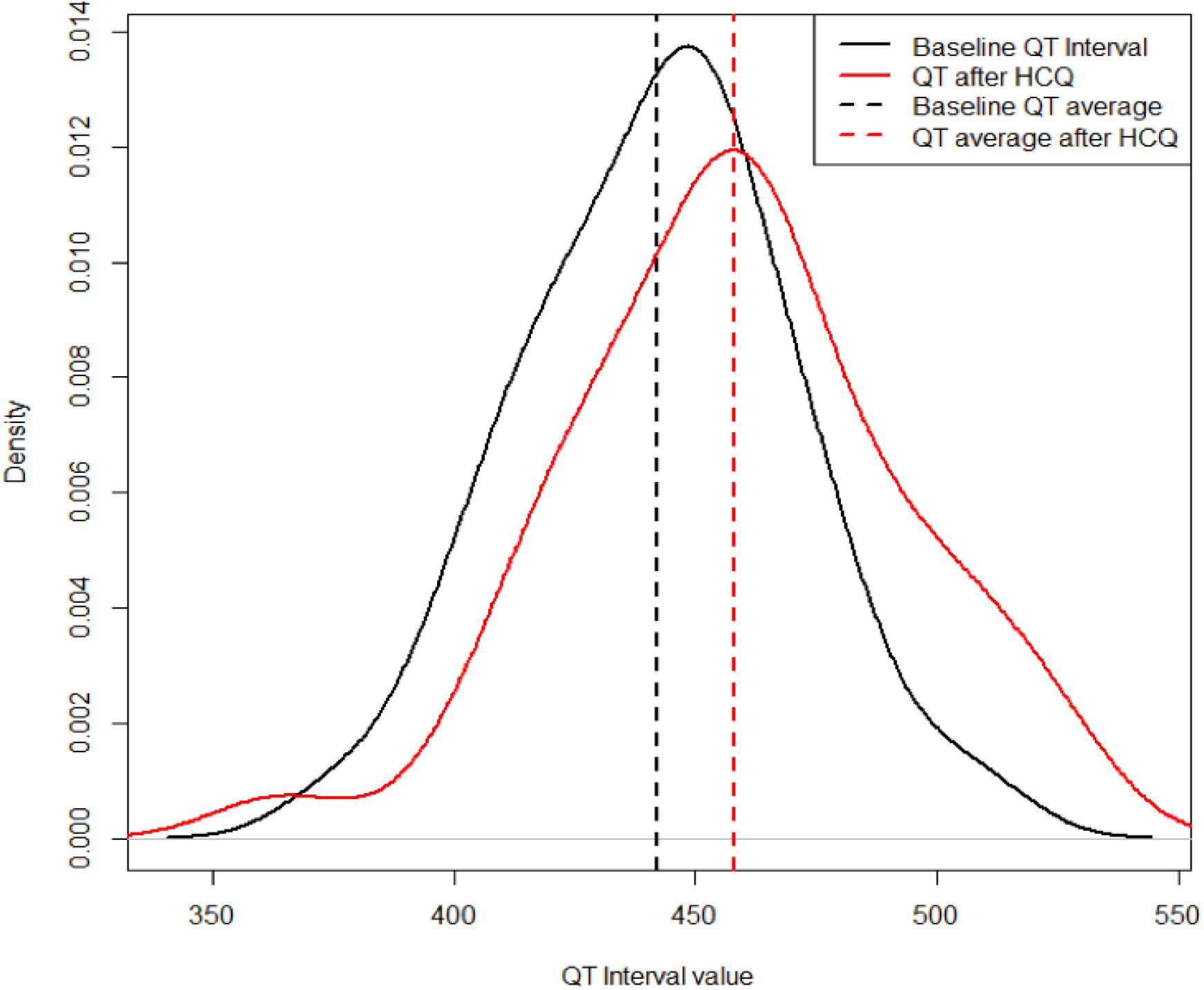
QT Interval Estimated Densities

**Figure 3.**
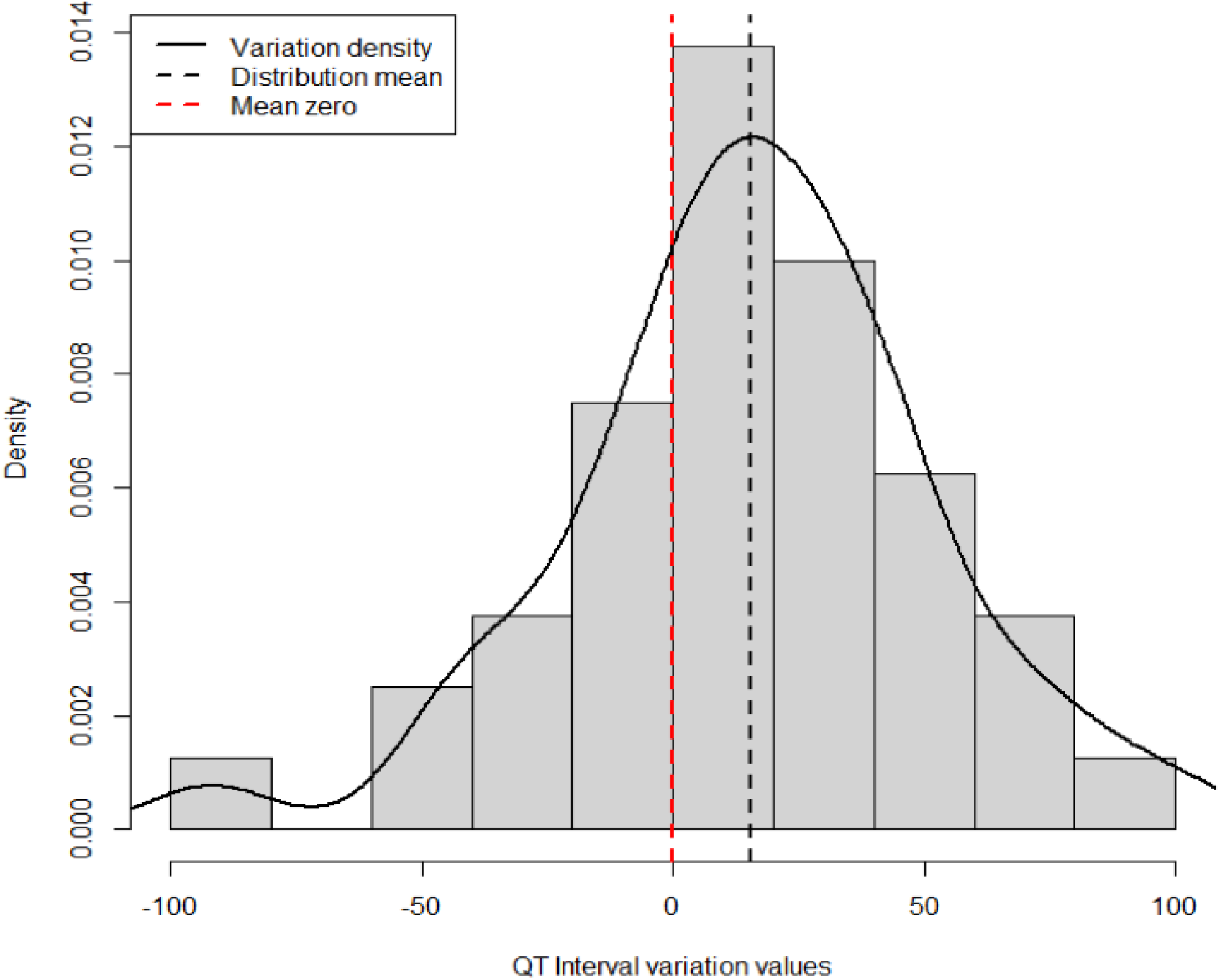
QT Interval Variation Distribution

Four patients (8.8%) experienced an interrupted treatment regimen. One because of significant prolongation in QTc, one for a non-disclosed reason, and two because of medical preference.

### Incidental Arrhythmias

No patient presented with TdP or any other ventricular polymorphic tachycardia during or after the treatment regimen. There were no arrhythmogenic deaths. As previously described, aborted cardiac arrests were non-arrhythmic (pulseless electrical activity or asystole).

Seven patients (15.5%) presented with arrhythmias during or immediately after the treatment regimen. One patient presented with frequent premature ventricular contractions (PVC) and ventricular bigeminy. Six patients (13.3%) presented with supraventricular tachycardia (SVT), two with atrial fibrillation, two with atrial flutter, and two showed paroxysmal supraventricular tachycardia. Most arrhythmias were hemodynamically stable and were treated with antiarrhythmic drugs. Atrial flutters were treated with synchronized electrical cardioversion.

As for conduction disturbances, one patient presented with right bundle branch block and one presented with left bundle branch block during the treatment regimen.

### Logistic Regression Model

In the univariate analysis, p values ≤0.10 were considered for multivariate analysis. The data are shown in Table 3. Age, sex, previous heart disease, and the use of QT prolonging medications were not significant predictors of a final QTc ≥480 ms. Comorbidities were separated into two groups: none/one, and two or more, and were also not significant predictors of a final QTc ≥480 ms. Azithromycin use, oxygen saturation at admission, acute heart failure, vasoactive drug use, and aborted cardiac arrest were also not significant predictors of a prolonged final QTc.

**Table 3.** Univariate Predictor Analysis

|  | *QTc < 480 ms* | *QTc ≥ 480 ms* | *p value* |
| --- | --- | --- | --- |
|  | 29 (72.5) | 11 (27.5) |  |
| *Age (y)* | 60.45 (18.57) | 66.36 (10.37) | 0.326 |
| *Gender (M)* | 18 (62.1) | 6 (54.5) | 0.942 |
| *Comorbidities ≤ 1* | 11 (37.9) | 3 (27.3) | 0.795 |
| *Comorbidities ≥ 2* | 18 (62.1) | 8 (72.7) | 0.795 |
| *Prolonging QTc Medications* | 11 (37.9) | 3 (27.3) | 0.416 |
| *Previous Heart Disease* | 6 (20.7) | 1 (9.1) | 0.692 |
| *Azithromycin* | 24 (82.8) | 9 (81.8) | 1.00 |
| *Basal QTc* | 437.72 (26.71) | 455.36 (29.07) | 0.076 |
| *O2 saturation (IQR)* | 0.88 (0.85-0.90) | 0.86 (0.84-0.88) | 0.190 |
| *Hemodialysis* | 11 (37.9) | 10 (90.9) | 0.008 |
| *Acute Heart Failure* | 1 (3.4) | 2 (18.2) | 0.364 |
| *Vasoactive Drugs* | 18 (62.1) | 10 (90.9) | 0.164 |
| *Aborted Cardiac Arrest* | 2 (6.9) | 1 (9.1) | 1.000 |
| *Troponin (IQR)* | 0.16 (0.16-0.16) | 0.20 (0.16-0.30) | 0.014 |
| *K (minimum value)* | 3.57 (0.51) | 3.18 (0.66) | 0.057 |
| *Mg (minimum value)* | 1.93 (0.23) | 1.72 (0.19) | 0.009 |
| *Creatinine (maximum value – IQR)* | 1.16 (1.00 – 1.70) | 2.07 (1.81 – 3.20) | 0.007 |

Reference values: Potassium (K), 3.5-5.5 mEq/L; magnesium (Mg), 1.6-2.6 mg/dL; troponin, <0.16 pg/mL; creatinine (Cr), 0.6-1.3 mg/dL. Continuous variables are expressed as mean ± standard deviation. Categorical variables are presented as n (%). IQR, interquartile range.

However, acute renal failure with increasing creatinine values and the requirement of renal substitution therapy with hemodialysis were statistically significant predictors of prolonged QTc, as were elevated troponin levels and low levels of potassium (K) and magnesium (Mg). The baseline QTc interval also reached statistical significance. These variables were then subjected to multivariate analysis utilizing logistic regression, which is presented in Table 4.

**Table 4.** Multivariate Predictor Analysis

| **Variable** | **OR** | **CI 95%** | **p-value** |
| --- | --- | --- | --- |
| Hemodialysis | 10.3494 | (1.0482, 102.1826) | 0.0455 |
| Creatinine | 1.8814 | (0.9721, 3.6412) | 0.0607 |
| Basal QTc | 0.9919 | (0.9247, 1.0641) | 0.8214 |
| K | 1.1809 | (0.1936, 7.2031) | 0.857 |
| Mg | 3.0049 | (0.2198, 41.0859) | 0.4096 |
| Troponin | 0.1434 | (0.0004, 58.693) | 0.5268 |

OR: odds ratio; CI: confidence interval.

The need for hemodialysis remained a significant predictor of QT prolongation ≥480 ms, even after adjustment for confounding factors, such as K and Mg values, azithromycin, comorbidities, and QTc-prolonging medications (OR, 10.34; 95% CI, 1.04–102; p = 0.045).

In addition, for each 1 mg/dL increase in creatinine levels, the risk for a final QTc ≥480 ms increased by 1.88 (95% CI, 0.97–3.6; p = 0.06). Although nearly significant, we think it is important to emphasize this data, as the clinical implications might be important, and the lack of significance is probably due to the small sample size. K, Mg, and troponin levels, as well as baseline QTc were not statistically significant predictors of final QTc ≥480 ms in multivariate analysis (Table 4).

## Discussion

This report demonstrates that a hemodialysis requirement is a strong predictor for QT interval prolongation in critically-ill patients with SARS-CoV-2 infection treated with HCQ. Acute renal failure and an increase in creatinine levels are possible predictors as well, as it neared significance, showing 1.88-fold risk for each 1 mg/dL increase in creatinine level. Concomitant QT prolonging medications and azithromycin association were not predictors for enlargement of the QT interval.

Although the arrhythmogenic risk of ventricular arrhythmias and TdP is markedly higher when QTc intervals are ≥500 ms, current reports and guidelines determine that QTc intervals longer than 480 ms are diagnostic of long QT syndrome, and should raise concern when monitoring drug-induced QTc.^8,9^ Moreover, Mercuro et al. reported a TdP episode in a patient with a QTc interval of 499 ms, treated with hydroxychloroquine.^10^ Thus, in this study, a cutoff of 480 ms was established for QT enlargement.

### HCQ Pharmacokinetics and Immunomodulatory Effects

Although used in clinical practice for almost 65 years, the 4-aminoquinolines antimalarial agents, chloroquine and hydroxychloroquine^11^ had never received the level of scrutiny as they have in the COVID-19 pandemic, since in vitro and anecdotal clinical trials showed antiviral activity against SARS-CoV-2.^1,2^

HCQ has immunomodulatory properties and has been used in rheumatologic diseases, such as systemic lupus erythematosus (SLE)^12^ and rheumatoid arthritis (RA).^13^ HCQ is a weak base, with complex pharmacokinetics and a large volume of distribution, metabolized in the liver in active metabolites.^11^ Urinary excretion is variable, accounting for 20–40% of the total elimination of the medication, including the unchanged drug and its active metabolites.^11,12^ Corroborating the findings in our trial, acidosis and impaired renal function are known clinical conditions that may increase the plasma concentration of the drug, leading to potential adverse effects, such as QT interval prolongation and TdP.^11^

### General Effects of HCQ in the Cardiovascular System

The cardiovascular effects of HCQ have been reported with divergent data. Liu et al published a meta-analysis involving 19 observational studies and 19,679 patients, showing a 30% reduction in cardiovascular risk, and reported favorable changes in the lipid profile.^14^ Mathieu et al. corroborated these findings with another meta-analysis involving 24,923 HCQ users and 36,327 non-users, demonstrating low diabetes incidence among HCQ users and a nearly significant decrease in the occurrence of cardiovascular events in the HCQ group.^15^ However, Chartre et al. showed, in a systematic review, cardiac complications with chloroquine and hydroxychloroquine use. Although reporting data from a reduced population (127 patients), with outcomes extracted from case reports and short series of cases and only 39.4% patients using HCQ, they demonstrated serious cardiovascular events associated with the chronic use of these drugs, such as conduction disorders occurring in 85% of patients, and heart failure in 26.8% of patients undergoing chronic treatment.^16^ This study showed no heart failure or other mechanical events attributable to HCQ, probably because of the short-term treatment. However, conduction disturbances were noted, such as right and left bundle branch block, even with such short-term use.

### HCQ, QT interval, and Arrhythmias

It is well known that chloroquine and HCQ prolong the QTc interval by blocking the inward K rectifier current (IKr), acting on the hERG channel.^17^ Besides promoting mild to moderate prolongation in QT interval, chloroquine and hydroxychloroquine have been linked to TdP.^17,18^

Nonetheless, reports in the literature are conflicting regarding the amount of QT interval enlargement and the incidence of ventricular polymorphic tachycardias, such as TdP, especially during the SARS-CoV-2 outbreak. Saleh et al. showed that the association of HCQ and azithromycin is safe for the treatment of patients hospitalized with SARS-CoV-2 infection, with no arrhythmic events, and despite a significant increase in the QT interval, discontinuation of the treatment regimen only occurred in 3.5% of patients.^19^ These findings are similar to those reported by Bun et al., who showed that in 73 patients hospitalized with SARS-CoV-2 infection, there were no drug-induced life-threatening arrhythmias, with a discontinuation rate due to QTc interval prolongation in 2.8% of patients.^20^ Cavalcanti et al. also showed that, in patients with mild to moderate SARS-CoV-2 infection, prolongation of the QTc interval ≥480 ms was more frequent in patients treated with HCQ, with or without azithromycin; however, there was no significant rise in arrhythmia incidence under this treatment regimen.^21^

However, Borba et al. showed that a high dosage of chloroquine (600 mg twice a day for 10 days) is associated with poor outcomes, with 18.9% patients a showing a QTc interval prolongation ≥500 ms and 2.7% experiencing ventricular tachycardia before death, but without a record of TdP.^22^ This is an important trial showing the effects of 4-aminoquinolines on the QT interval. In our trial, patients were treated with HCQ, known for carrying less arrhythmogenic effects than chloroquine,^16^ and administered in a much lower dosage. Nonetheless, the use of different doses of chloroquine as well as the clinical suspicion, not confirmation, of SARS-CoV-2 infection used as inclusion criteria, limit the capability of this trial for comparison.

Regarding proper HCQ use and arrhythmic events, Chorin et al. reported, in 251 inpatients with SARS-CoV-2 infection, a high incidence of QT interval prolongation (23%), with basal QTc = 439 ± 29 ms to 473 ± 36 ms on day 4 of the treatment regimen with HCQ plus azithromycin. One patient (0.4%) presented with TdP and required electrical cardioversion.^23^ Confirming the findings in our trial, renal function worsened by 33% on the day of maximum QT interval measurement.^23^ Moreover, as previously described, Mercuro’s trial reported a TdP episode in a patient with a QTc interval of 499 ms who had been treated with hydroxychloroquine.^10^

### HCQ and Predictors for QT Enlargement and TdP

Our findings are similar to the report by Hooks et al., who showed that in 819 patients treated for rheumatologic diseases, HCQ use is associated with QT interval prolongation, and a significant risk of prolongation is related to a clinical history of chronic kidney disease (CKD). They also reported that atrial fibrillation and heart failure are risk factors for QT enlargement.^24^

Maraj et al. reported similar rates of QT prolongation (23%) compared with our trial (27.5%) among inpatients treated with HCQ plus azithromycin for SARS-CoV-2 infection.^25^ This study showed that 14% of patients had a QTc interval ≥500 ms, and there was one case of TdP. Among other risk factors, this report demonstrates that 43% of patients presenting with significant QTc interval prolongation had baseline renal insufficiency or developed acute renal failure, corroborating the findings in our trial.^25^

Interestingly, Tsidale and colleagues developed a risk score to predict QT interval prolongation in hospitalized patients that does not contemplate renal dysfunction as predictor of QT enlargement.^26^ However, regarding drugs with moderate to significant renal excretion, this might be a decisive parameter to consider. Drawing attention to this matter, the 2010 Scientific Statement of the American Heart Association for the prevention of TdP in hospital settings addresses the burden of renal dysfunction as a risk factor for TdP, advising clinicians to monitor the QTc interval and electrolyte variations closely, especially in patients receiving drugs with long half-lives and substantial renal excretion.^27^ The findings of this study are in accordance with such recommendations.

### Current HCQ Use for Covid-19 and Other Possible Employments in Critical Patients

Several reports did not show improvement in clinical status under chloroquine or HCQ treatment for patients with SARS-CoV-2 infection^21,22,28^. Consequently, these drugs are not approved as first line therapy for SARS-CoV-2 infection in Brazil anymore.

Despite that, it is important to recognize the arrhythmogenic effects of such treatment in this subset of patients, as well as its predictors, to prevent rare but catastrophic life-threatening arrhythmias. Furthermore, HCQ might be utilized for other critical conditions such as SLE patients with end-stage renal disease^29^, lupus nephritis^30^, as well as anti-cancer therapies^31,32^. Clinicians treating this conditions must be aware that patients presenting with renal dysfunction and needing substitutive renal therapy with hemodialysis are at risk for QT interval enlargement and its associated arrhythmias.

Although previous trials described more frequent prolongation of QT interval during renal dysfunction, hemodialysis need in the critically-ill patient has not been reported as an independent predictor of QT interval enlargement yet. Therefore, we believe it is important to describe and emphasize that worsening in renal function requiring substitutive renal therapy with hemodialysis is an independent predictor for QT interval enlargement under HCQ treatment.

### Limitations

Our study has several limitations. First, this was a retrospective observational trial, with a non-randomized sample of patients. The treatment was not homogeneous between the patients, and 17.8% of patients were treated only with HCQ without azithromycin.

Moreover, the sample size, although large enough to demonstrate hemodialysis as a predictor of QTc lengthening, was not large enough to show that acute renal failure and elevated creatinine levels are also statistically significant predictors, despite the fact that it can be inferred, reaching nearly significance.

Furthermore, the inability to measure the final QTc interval in five patients might also affect the outcomes, given the reduced sample size. The ECG review by only one electrophysiologist might also have contributed to measuring bias. Lastly, we did not assess serum drug concentrations.

## Conclusions

Despite great controversy, HCQ has been used in a large number of patients in critically ill clinical conditions with SARS-CoV-2 infection. Knowledge of the pharmacological effects of this drug in this subset of critically-ill patients is very important not only in the case of COVID-19 treatment, but also in other critically ill clinical conditions, such as SLE patients with end-stage renal disease, lupus nephritis and severe cases of malaria.

This trial demonstrates that HCQ promotes mild to moderate lengthening in the QT interval in most cases. However, patients with renal function impairment and those evolving with the need for substitution renal therapy with hemodialysis are at risk for serious QT prolongation, and should be monitored more closely. Avoidance of electrolyte disturbances and careful continuous measurement of the QT interval are crucial.

Despite these findings, larger, randomized clinical trials are needed to establish the safety and efficacy of HCQ in the setting of SARS-CoV-2 infection.

## Data Availability

The authors confirm that the data supporting the findings of this study are available within the article [and/or] its supplementary materials.

## Acknowledgements

We thank all the clinical and allied professional teams involved in the care of patients with SARS-CoV-2 infection.

